# The epidemiologic parameters for COVID-19: A Systematic Review and Meta-Analysis

**DOI:** 10.1101/2020.05.02.20088385

**Authors:** Neda Izadi, Niloufar Taherpour, Yaser Mokhayeri, Sahar Sotoodeh Ghorbani, Khaled Rahmani, Seyed Saeed Hashemi Nazari

## Abstract

**Introduction:** The World Health Organization (WHO) declared the outbreak to be a public health emergency and international concern and recognized it as a pandemic. The aim of this study was to estimate the epidemiologic parameters of novel coronavirus (COVID-19) pandemic for clinical and epidemiological help.

**Methods:** Four electronic databases including Web of Science, Medline (PubMed), Scopus and Google Scholar were searched for literature published from early December 2019 up to 23 March 2020. The “*metan*” command was used to perform a fixed or random effects analysis. Cumulative meta-analysis was performed using the “*metacum*” command.

**Results:** Totally 76 observational studies were included in the analysis. The pooled estimate for R_0_ was 2.99 (95% CI: 2.71-3.27) for COVID-19. The overall R_0_ was 3.23, 1.19, 3.6 and 2.35 for China, Singapore, Iran and Japan, respectively. The overall Serial Interval, doubling time, incubation period were 4.45, 4.14 and 4.24 days for COVID-19. In addition, the overall estimation for growth rate and case fatality rate for COVID-19 were 0.38% and 3.29%, respectively.

**Conclusion:** Calculating the pooled estimate of the epidemiological parameters of COVID-19 as an emerging disease, could reveal epidemiological features of the disease that consequently pave the way for health policy makers to think more about control strategies.

## Introduction

Coronaviruses are a group of RNA viruses that cause diseases among humans and animals (1). The latest of coronavirus types as a novel coronavirus that was named severe acute respiratory syndrome coronavirus 2 (SARS-Cov2) or COVID-19 occurred in Wuhan, China in December 2019 with a human outbreak (2).

The World Health Organization (WHO) declared the outbreak to be a public health emergency and international concern and recognized it as a pandemic on 11 March 2020 (3). COVID-19 has widely spread in the world and is prevalent in different countries such as China, Italy, United states, France, Spain, Iran and Germany with 2,833,697 cases and 197,354 deaths and 807,469 recovered until 24 April in the whole world (4). The main rout of transmission of COVID-19 is based on human-to-human transmission via either respiratory droplets, saliva or close contacts with infected people or aerosol generation procedures during clinical care of COVID-19 patients (5).

Most COVID-19 infected people (80.9%) are with mild to moderate respiratory syndromes, old people or patients with underlying diseases such as diabetes, cardiovascular disease, cancer, immune deficiency and respiratory disease are more at risk to develop sever (13.8%) and critical (4.7%) disease (6,7).

Knowledge regarding epidemiological characteristics and parameters of the infectious diseases such as, incubation period (time from exposure to the agent until the first symptoms develop), serial interval (duration between symptom onset of a primary case and symptom onset of its secondary cases), basic reproduction number (R_0_) (the transmission potential of a disease) and other epidemiologic parameters is important for modelling and estimation of epidemic trends and also implementation and evaluation of preventive procedures (8–11).

About COVID-19 pandemic parameters, there are many reports from different countries in the world. For example, about 25.6 % to 51.7% of patients have been reported to be asymptomatic or with mild symptoms (12) and 25-30% of them have been admitted to ICU for medical care (13). Case-fatality rate was reported in China and other countries among old patients 6% (4-11% ranges) and 2.3 % in all ages (13,14). Furthermore, the median incubation period was reported as 5-6 days (2-14 ranges) from WHO while, in China incubation period was reported up to 24 days (15,16). Also, according to the different mathematical models, R_0_ was reported about 6.47 (1.66-10 ranges) in China, 2.6 in South-Korea and 4.7 in Iran (17,18,19).

Thus, according to the reports from different countries about epidemiological characteristics of COVID-19 pandemic, different methods and different values of parameters have been observed. So, for efficient estimation and forecasting of disease spreading, we need acceptable and real values of each parameter. The present study was conducted to provide a systematic assessment and estimation of parameters related to COVID-19. This evaluation will help researchers with better prediction and estimation of current epidemic trends.

## Method

The current study is a systematic review and meta-analysis to determine the epidemiologic parameters for COVID-19.

### Search Strategy

To find relevant studies, a comprehensive literature search of the Web of Science, Medline (Pubmed), Scopus and Google Scholar was performed for observational studies published electronically from early December 2019 up to 23 March 2020.

Two researchers independently searched studies. In search strategy, English keywords and probable combination of them were used. The keywords included “novel coronavirus” ، “2019-noCov”، “COVID-19”، “basic reproduction number”، “serial interval”̜ “incubation period” “،doubling time”، “growth rate”̜ “case-fatality rate” ̜“mortality rate”، “onset of symptom to hospitalization”. The Boolean operators (‘OR ‘and ‘AND’) were used for combination of keywords. The search strategy was as follows:

**Keywords:** (novel coronavirus OR 2019-nCov OR COVID-19) AND (basic reproduction number OR basic reproductive rate) OR (case fatality rate OR case fatality ratio) OR mortality rate OR doubling time OR growth rate OR incubation period OR onset of symptom to hospitalization.

### Study Selection

Consistent with PRISMA guidelines, the standard meta-analysis techniques, we included studies. All of the extracted articles independently were screened by two researchers. Abstract and full text of the articles were reviewed and duplicated studies were excluded and then relevant articles were selected for data extraction.

### Study inclusion and exclusion criteria

All epidemiological studies designs (observational studies) including peer reviewed or not peer reviewed articles that provided the epidemiologic parameters of interest regarding the novel corona virus were included. In addition, irrelevant studies, letters and news and studies that didn’t report epidemiologic parameters were excluded.

### Screening and Data extraction

All articles were reviewed independently by four researchers and information was extracted using designed checklist (**Appendix 1**). Extracted items were name of the first author, years and month of article published, duration of the study, location of study conduction, type of parameters, point estimate or mean/median and its confidence interval for epidemiological parameters and review status of articles (peer-reviewed or not).

### Quality assessment of studies

To assess the quality of included the peer-reviewed and not peer reviewed articles, we used the STROBE quality assessment scale for observational studies. We assessed the quality of all studies and finally, studies with high and medium quality were included in the analyses.

### Statistical analysis

The “metan” command was used to apply a fixed or random effects model based on Cochran’s Q-test results or a large Higgins and Thompson’s I^2^ value. Forest plots were used for graphical description of the results. Also, the “metacum” command was used for cumulative meta-analysis to determine trend of basic reproductive number (R_0_).

In studies that mortality rate was reported, because of the denominator was confirmed cases, it was considered a case fatality rate (CFR). In addition, for studies that reported the median and interquartile range (IQR), the median was considered equivalent to the mean and the IQR was converted to standard deviation using the “IQR/1.35” formula. Stata 14 was used for all statistical analyses.

## Results

Having assessed the quality of relevant studies, 76 observational studies up to 23 March, 2020 were included in this study (**Follow diagram**). The majority of studies were done in Wuhan, China. Detailed information of the eligible studies and their characteristics has been shown in **Appendix 1** (12,17,18,20–92).

- **The overall basic reproductive number (**R_0_**) by country and peer review status** **Total:** The overall R_0_ was 2.99 (95% CI: 2.71-3.27) for COVID-19 (**Table 1**). **Country:** The overall R_0_ was 3.23, 1.19, 3.6 and 2.35 for China, Singapore, Iran and Japan, respectively (**Table 1**). **Peer review status:** The overall R_0_ was 2.75 and 3.08 for peer reviewed and not peer reviewed articles, respectively (**Table 1**).
- **The overall serial interval (SI) by country and peer review status** **Total:** The overall SI was 4.45 days (95% CI: 4.03-4.87) for COVID-19. **Country:** Using random effect model, the overall SI was 4.46 and 4.64 days for China and Singapore, respectively (**Error! Reference source not found**.). **Peer review status:** The overall SI was 5.3 and 4.39 days for peer reviewed and not peer reviewed articles, respectively (**Error! Reference source not found.2**).
- **The overall doubling time by peer review status** **Total:** The overall doubling time was 4.14 days (95% CI: 2.67-5.62) for COVID-19. **Peer review status:** The overall doubling time was 3.33 and 4.64 days for peer reviewed and not peer reviewed articles, respectively (**Error! Reference source not found.3**).
- **The overall incubation period by peer review status** **Total:** The overall incubation period was 4.24 days (95% CI: 3.03-5.44) for COVID-19. **Peer review status:** The overall incubation period was 4.03 and 5.82 days for peer reviewed and not peer reviewed articles, respectively (**Table 1**).
- **The overall estimation for other epidemiologic parameters** The overall estimation for growth rate and case fatality rate for COVID-19 were 0.38% and 3.29%, respectively (**Table 1 & Fig 4**). In addition, the overall time from symptom onset to hospitalization was 5.09 days for COVID-19 (**Table 1**).
- **The trend of** R_0_ **for COVID-19** Based on the cumulative meta-analysis, the trend of R_0_ had been increasing at first and, then, decreasing in March.

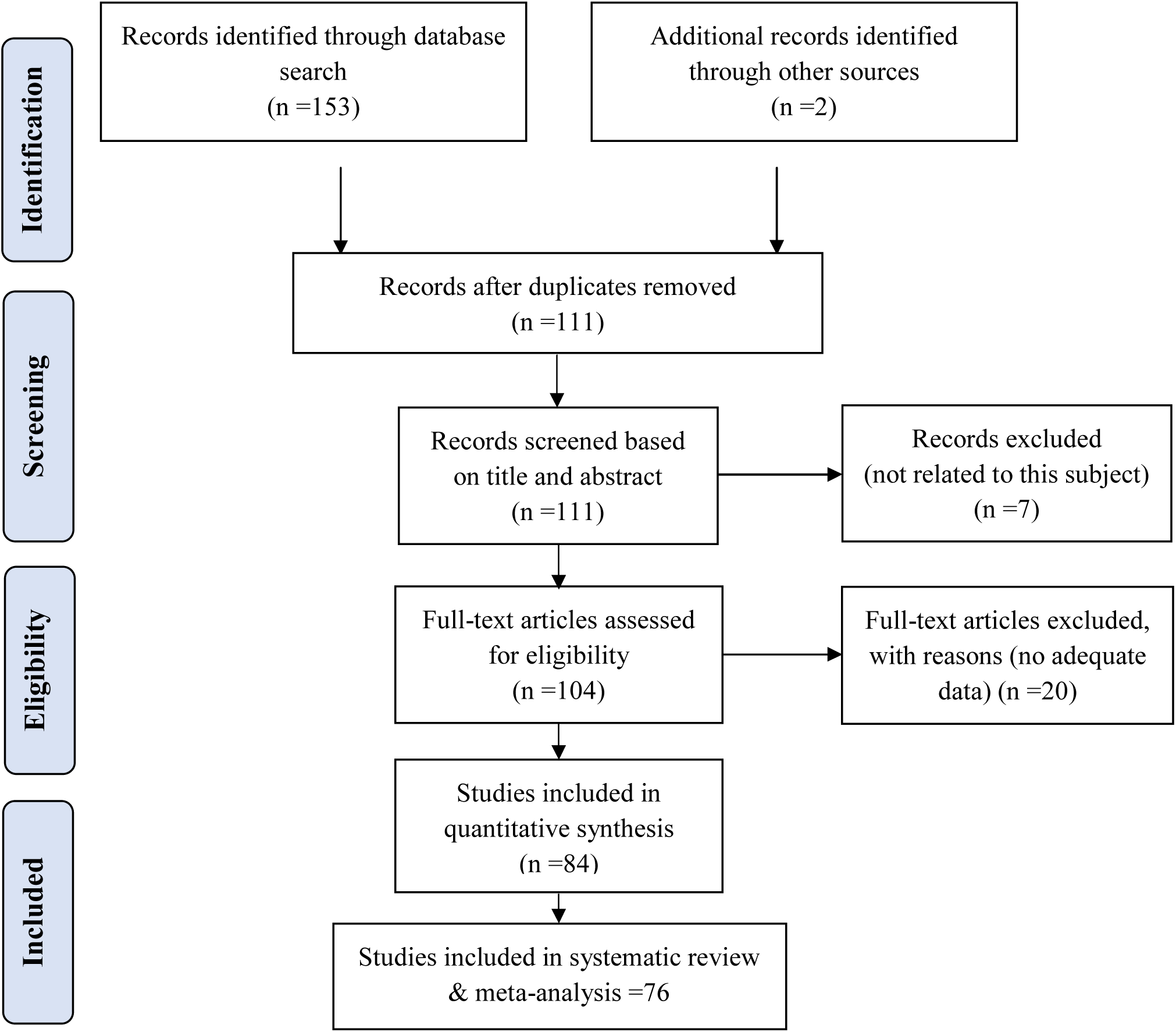
**Flow diagram of the study selection process and including publications for the epidemiologic parameters for COVID-19**

**Figure 1.**
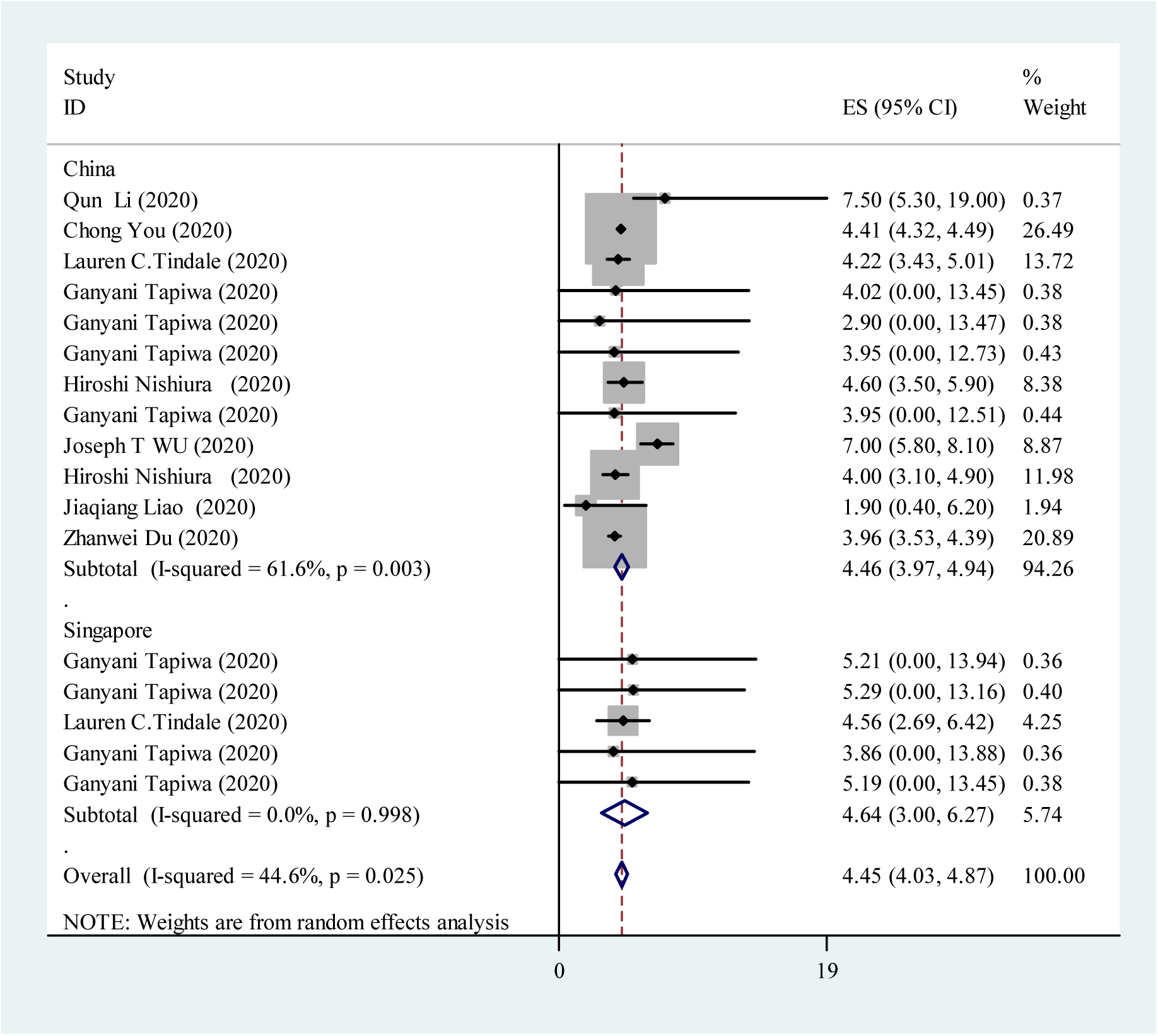
The overall serial interval (SI) for COVID-19 by country.

**Figure 2.**
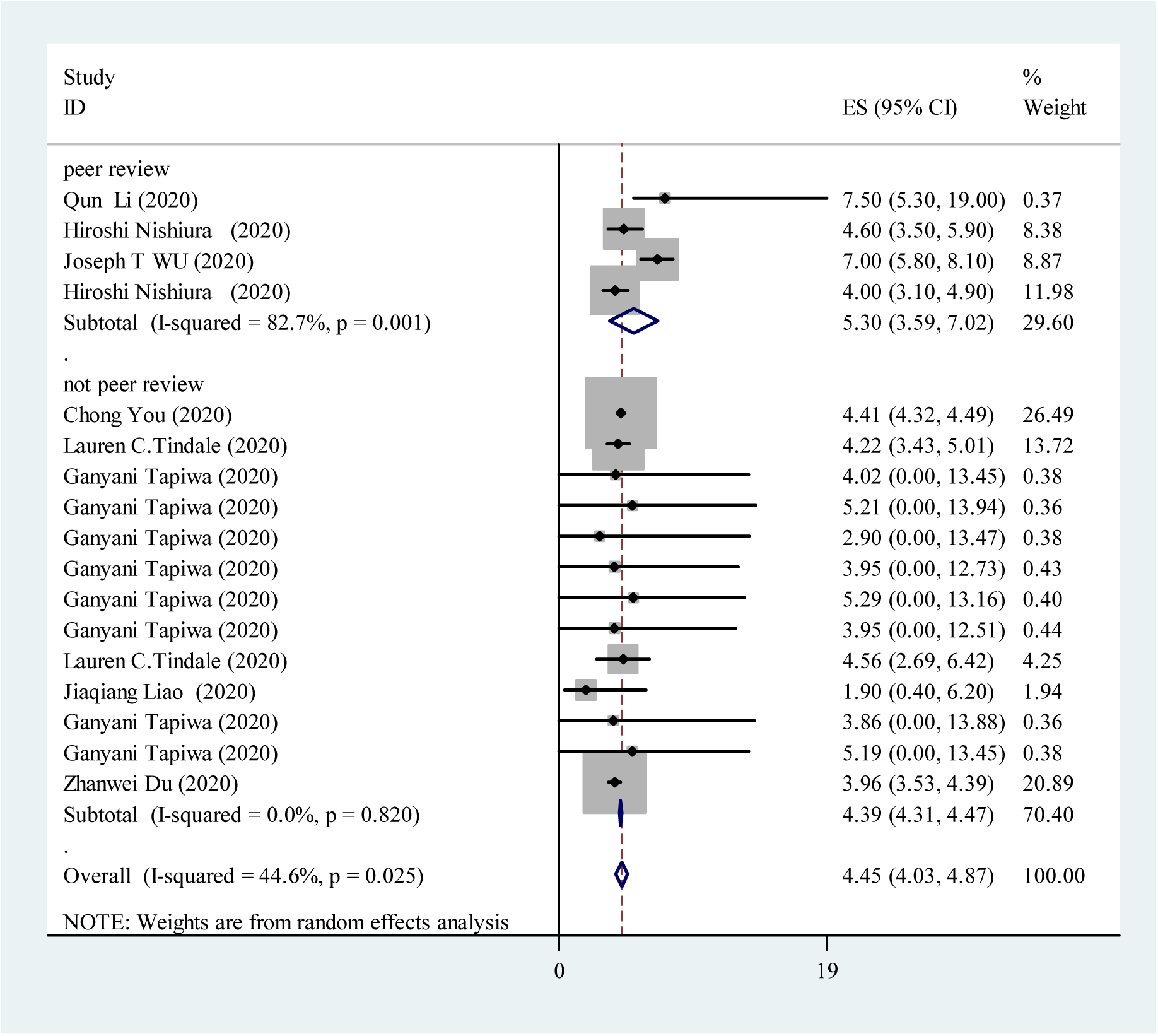
The overall serial interval (SI) for COVID-19 by peer review status.

**Figure 3.**
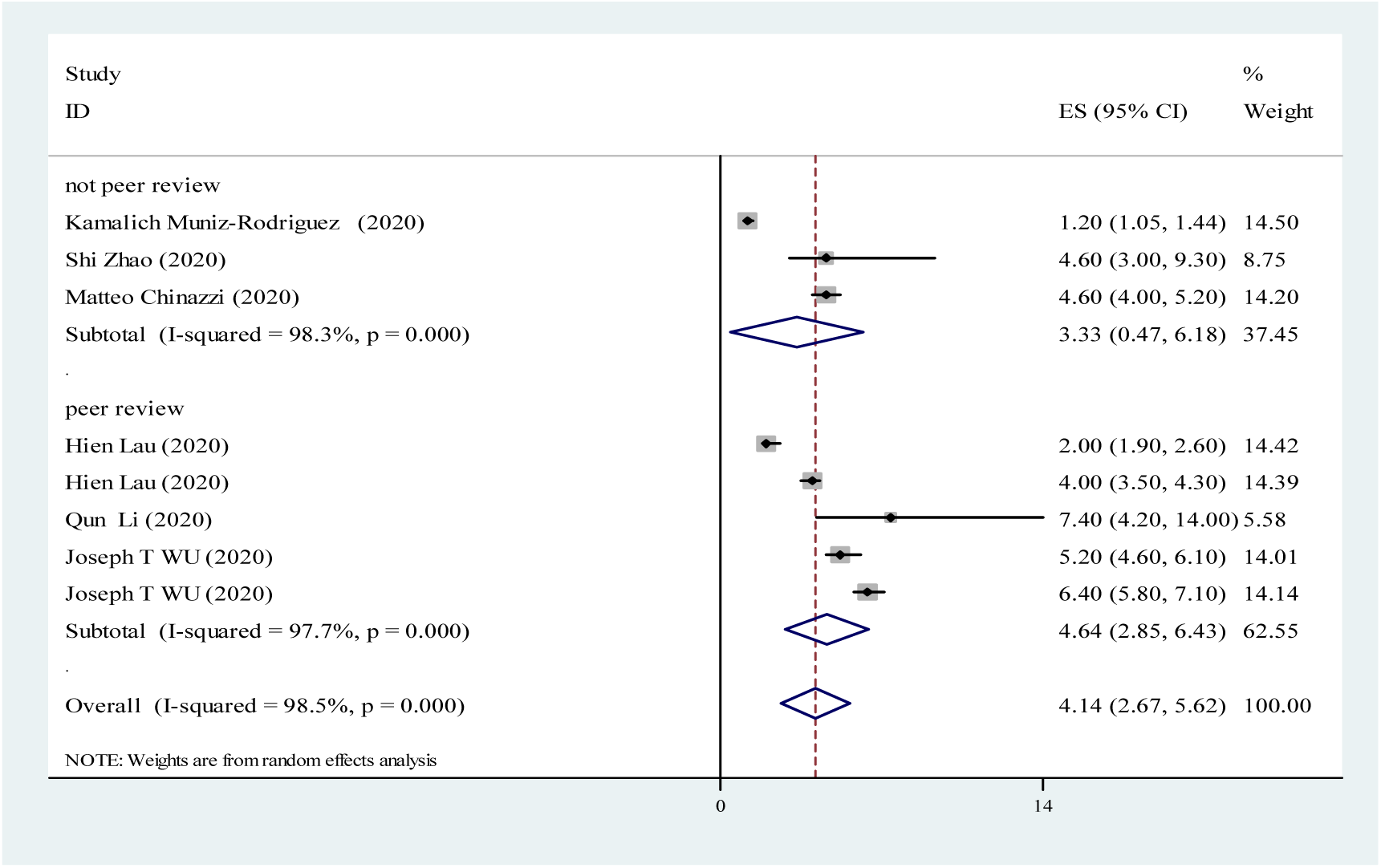
The overall doubling time for COVID-19 by peer review status.

**Figure 4.**
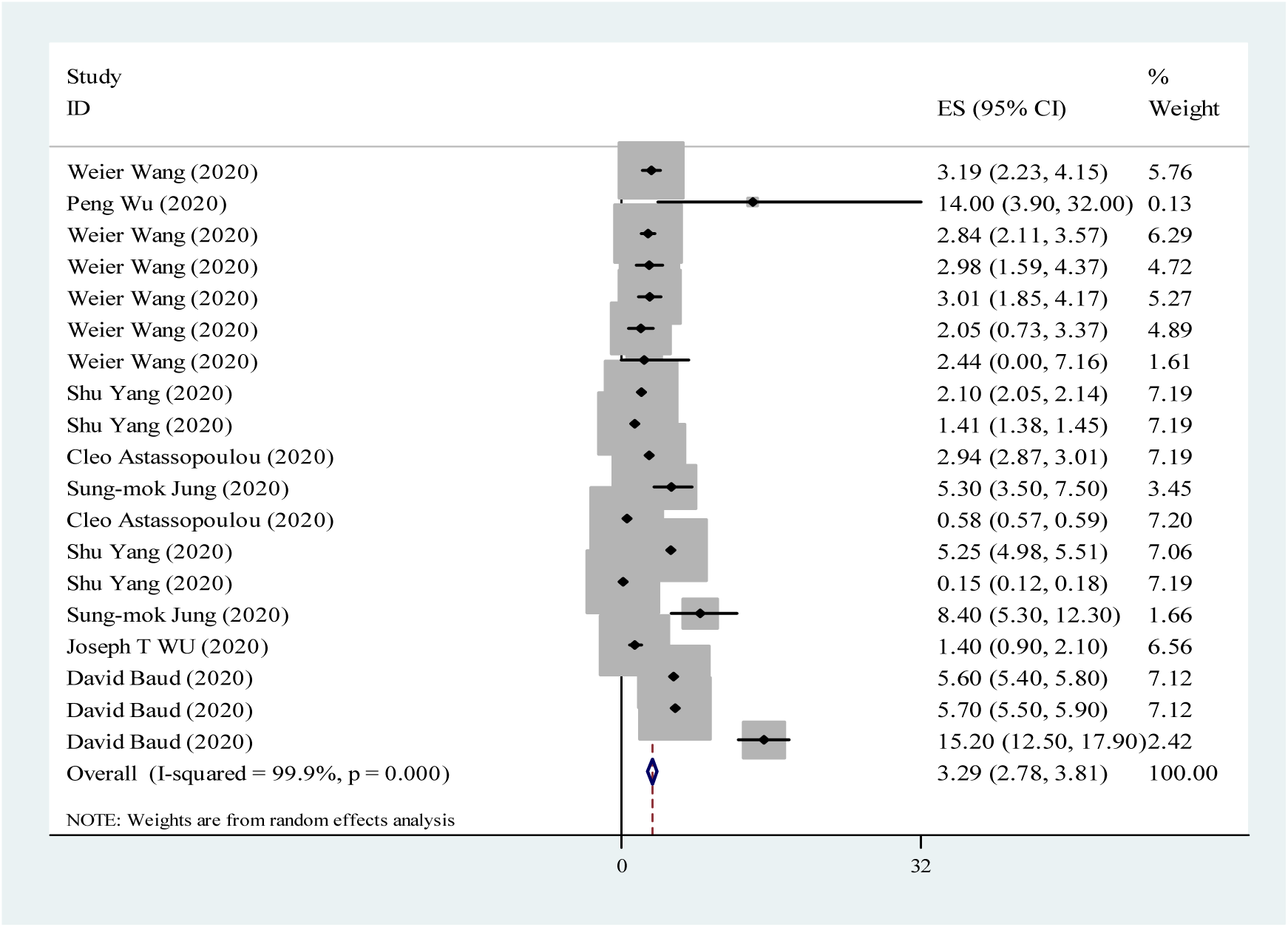
The overall case fatality rate (CRF) for COVID-19.

**Table 1.** The overall estimation of epidemiologic parameters for COVID-19.

| Parameters |  | No. of studies | Estimate | 95% CI | P for Heterogeneity | I<sup>2</sup> (%) |
| --- | --- | --- | --- | --- | --- | --- |
| **Basic Reproductive Number (R<sub>0</sub>)** | **Overall** | 69 | 2.99 | 2.71-3.27 | <0.001 | 99.3 |
|  | **Korea** | 1 | 2.6 | 2.5-2.7 | - | - |
|  | **China** | 57 | 3.23 | 2.92-3.55 | <0.001 | 99.1 |
|  | **Singapore** | 6 | 1.19 | 1.07-1.3 | <0.001 | 82.2 |
|  | **Iran** | 2 | 3.6<sup>a</sup> | 3.1-4.09 | 0.99 | - |
|  | **Japan** | 3 | 2.35 | 2.1-2.6 | 0.007 | 80.1 |
|  | **Peer Review** | 13 | 2.75 | 2.25-3.24 | <0.001 | 99.4 |
|  | **Not Peer Review** | 56 | 3.08 | 2.73-3.43 | <0.001 | 99.3 |
| **Growth Rate (%)** | **Overall** | 5 | 0.38 | 0.2-0.55 | <0.001 | 97.7 |
| **Symptom onset to Hospitalization (day)** | **Overall** | 6 | 5.09 | 2.15-8.02 | 0.03 | 53 |
| **Incubation Period (day)** | **Overall** | 22 | 4.24 | 3.03-5.44 | 0.02 | 35 |
|  | **Peer Review** | 18 | 4.03 | 2.72-5.33 | 0.01 | 41 |
|  | **Not Peer Review** | 4 | 5.82<sup>a</sup> | 2.91-8.74 | 0.76 | 16 |

<sup>a</sup> Fixed effect model

## Discussion

In this secondary analysis, we aimed to calculate the pooled estimate of some epidemiological parameters of COVID-19 namely basic reproductive number (R_0_), serial interval, doubling time, incubation period, growth rate, case fatality rate (CFR), and time from symptom onset to hospitalization. Overall, the estimates were 2.99, 4.45 days, 4.14 days, 4.24 days, 0.38%, 3.29%, and 5.09 days, in the same order. Considering urgent issues in pandemic situation, peers have not reviewed some published papers. Therefore, we tried to calculate the pooled parameters by peer review status.

It should be noted that, R_0_ variations to some extent might be due to different methods calculations including exponential growth method, maximum likelihood, and Bayesian time-dependent method (93–95).

According to our results the pooled estimate of CFR 3.29% (95% CI: 2.78-3.81) is lower than SARS-CoV(96) and MERS-CoV (97). Health control policies, medical standard, and detection rate could affect CFR (35). Moreover, CFR estimate in the early phase of the epidemic might be biased (overestimated). Usually in the early phase, some subclinical cases and patients with mild symptoms may not be detected (detection bias) (98,99).

Pooled estimate of incubation period using 22 studies was 4.24 days (95% CI: 3.03, 5.44). Valid and precise estimate of incubation period has a pivotal role for duration of quarantine (50). In fact, knowledge about incubation period is useful for surveillance and control approaches, also modeling and monitoring activities (100).

Our estimate for overall doubling time-time for a given quantity to double in size or number at a constant growth rate was 4.14 days (95% CI: 2.67, 5.62). The doubling time has an important implication for predicting epidemic. Generally, social distancing, quarantine, and active surveillance are needed to reduce transmission and consequently extend the doubling time (101). Moreover, the authors tried to estimate pooled measures for growth rate and serial interval. These two epidemiological parameters are used to estimate reproduction number (102).

As a limitation, all 76 studies (except for one, Mirjam E Kretzschmar et al) (103) have been conducted in Asia, particularly in Wuhan, China. Some epidemiological parameters in Europe, Africa, and America could be different based on control strategies. Hence, distribution of these epidemiological parameters could be more globally. Future studies to calculate more generalized pooled estimates, using studies all over the world, would be recommended.

## Conclusions

Calculating the pooled estimate of the epidemiological parameters of COVID-19 as an emerging disease, could reveal epidemiological features of the disease that consequently pave the way for health policy makers to think more about control strategies.

## Data Availability

The data of this study are available in appendix file

## Acknowledgment

We would like to appreciate all those researchers who helped us to conduct this study.

